# Chronic Low Back Pain Patient Satisfaction with Lumbar Steroid Injection: a Data-Driven Analysis

**DOI:** 10.1101/2025.04.10.25325570

**Authors:** Maria Monzon, Iara De Schoenmacker, Andrea Cina, Reka Enz, Christian Lanz, Fabio Galbusera, Catherine R. Jutzeler, Zina-Mary Manjaly

**Affiliations:** ETH Zurich, Department of Health Sciences and Technology (DHEST), Zü rich, 8092, Switzerland; Swiss Institute of Bioinformatics (SIB), Lausanne, 1015, Switzerland; Schulthess Clinic, Department of Teaching, Research and Development, Zü rich, 8008, Switzerland; Schulthess Clinic, Department of Neurology, Zü rich, 8008 Switzerland

## Abstract

Chronic low back pain (CLBP) is a prevalent condition significantly reducing quality of life. Lumbar steroid injections are a widely used conservative treatment option, but their effectiveness varies among patients. This study aimed to develop a predictive framework that integrates clinical variables and patient demographics to evaluate post-treatment pain satisfaction in CLBP patients undergoing lumbar injection therapy. We performed a retrospective analysis of 212 CLBP patients to evaluate the treatment satisfaction and pain intensity changes using the Numerical Rating Scale (NRS). A Random Forest model, validated through nested cross-validation, achieved an average precision of 0.865 in predicting treatment satisfaction. SHapley Additive exPlanations (SHAP) analysis revealed pain self-efficacy features, particularly coping mechanisms and household activities, as key outcome predictors of post-treatment pain satisfaction. Clinically significant pain reduction thresholds were identified at an absolute change of 2.09 and a relative change of 30 % on the NRS. Our findings reveal the biological and social factors influencing post-treatment pain in CLBP patients. The identified pain reduction thresholds and predictors may help clinicians to develop individualized management strategies, optimizing treatment outcomes and improving patient care. Future research should refine the predictive model by incorporating additional multimodal variables to better capture CLBP heterogeneity.

## Introduction

Low back pain (LBP) is a highly prevalent condition that affects more than 600 million people worldwide^1^, with a lifetime prevalence of up to 80%^2–4^. Approximately 10% of the cases persist for over 3 months, meeting the criteria for chronic low back pain (CLBP). The rising prevalence of CLBP^5^ poses significant challenges for individuals and public health systems^6^, as it remains the leading cause of years lived with disability worldwide^1,7^. Low back pain, defined as discomfort between the costal margins and the inferior gluteal folds, is often accompanied by leg pain and may present with additional symptoms such as stiffness, reduced range of motion, muscle spasms, localized tenderness, paresis, numbness, or tingling^8^. It can result from various causes, including nerve injury, spinal cord compression, muscle or ligament damage, inflammation, or infection^8,9^. The location and characteristics of pain can provide clues to its etiology, but identifying the precise source and determining the optimal treatment in clinical practice often requires considerable trial and error. This process is further complicated by the fact that the etiology of CLBP is frequently linked to psychosocial factors, with patients commonly reporting symptoms such as poor concentration, disrupted sleep, memory difficulties, and irritability^10^. The biopsychosocial model of pain^11^ describes the complex interplay among biological, psychological, and social factors contributing to the pathophysiological heterogeneity of CLBP. This complexity likely underlies and explains the considerable variability in treatment effectiveness^12^.

Lumbar injection, which involve administering local anesthetics and steroids to structures of the lumbar spine^13^, is a therapeutic option for patients with CLBP who do not respond to first-line analgesics and physical therapy. Despite its widespread use, evidence regarding the effectiveness of infiltration therapy for CLBP remains inconsistent, with studies reporting mixed outcomes across patients^14,15^. While some patients experience significant symptom relief, others gain little or no benefit, possibly due to differences in pain perception, psychological factors, and comorbidities^16,17^. Recognizing this variability, previous research emphasizes the need to understand the mechanisms driving treatment outcomes and develop individualized treatment strategies^17–19^. Personalized care approaches, using prognostic profiling and clinical prediction models, have demonstrated potential to improve treatment outcomes^20,21^. Although predictive tools show potential in identifying patients who could benefit from infiltration therapy^22^, their clinical implementation and utility remain very limited^23^. A significant barrier to progress is the absence of a well-defined outcome measure tailored specifically to assess the success of lumbar injection therapy^24^.

Currently, the severity of CLBP and its associated disability are commonly assessed using patient-reported outcome measures (PROMs)^22,25,26^, such as the numerical rating scale (NRS)^27^ and visual analog scale (VAS)^27^ for pain intensity. Establishing a threshold that represents the smallest change in PROM scores perceived as beneficial by patients is essential to determine the clinical significance of a therapy^28,29^. While previous studies have explored such thresholds in surgical contexts^30–33^, there is a lack of studies addressing this need for lumbar steroid injection therapy. To bridge this gap, we leveraged data-driven methodologies to develop a comprehensive predictive framework for lumbar injection therapy in patients with CLBP by integrating clinical data and patient-specific demographics. First, we developed a predictive model to identify key factors influencing the effectiveness of lumbar steroid injection therapy, enabling the identification of patients most likely to experience improvements in pain perception. Treatment success was evaluated based on self-reported pain satisfaction following therapy. Next, we aimed to establish clinically relevant thresholds for pain reduction specific to infiltration therapy, focusing on the minimal reduction in pain scores required for patients to perceive treatment outcomes as satisfactory.

## Methods

### Study Design and Participants

A retrospective secondary analysis was performed using data from the *Treatment Expectation and their Influence on Infiltration outcome* (TREXI) study^16^. The TREXI study was a prospective observational longitudinal investigation carried out between February 2019 and December 2020 at the Department of Neurology, Schulthess Clinic in Zurich, Switzerland. The original cohort included 306 adult patients, aged 18 to 93 years, diagnosed with CLBP. For this secondary analysis, a subset of 212 patients who provided informed consent for the additional use of their data in research was included (Fig. 1a). The study was approved by the Cantonal Ethics Committee of Zurich (BASEC-NR 2023-02210) and complied with the ethical principles outlined in the Declaration of Helsinki.

**Figure 1.**
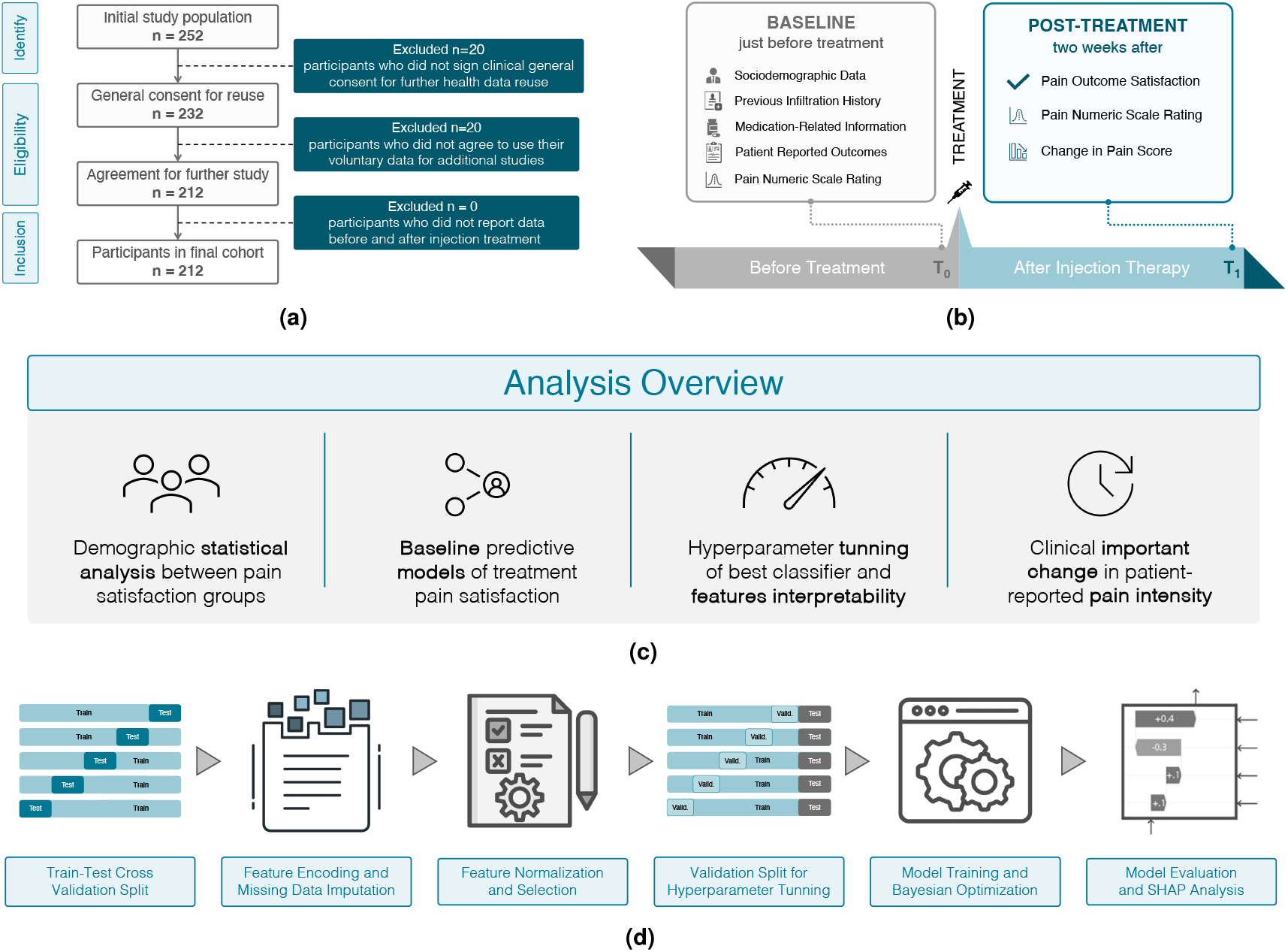
Study Design for Predicting Patient Satisfaction with Lumbar Steroid Injection Therapy. **(a) Retrospective cohort selection**: The flowchart illustrates the inclusion procedure of participants for the retrospective cohort analysis. **(b) Data collection timeline** for evaluating lumbar steroid injection outcomes in chronic low back pain patients, including baseline assessments (*T*_0_) and two-week post-treatment follow-up (*T*_1_). **(c) Statistical Analysis framework** comprising demographic variable analysis and machine learning baseline predictive modeling, classifier optimization incorporating feature selection, model hyperparameter tuning and feature importance (SHAP) analysis, and ROC curve analyses for clinical significance of minimal change in reported pain metrics. **(d) Predictive model development:** The process starts with nested cross validation, splitting data for model evaluation and hyperparameter tuning. Data preprocessing includes managing missing values, encoding categorical variables, and scaling features. Feature engineering includes feature normalization and selection, where the most informative ones are chosen based on statistical tests. Model tuning in the inner loop optimizes the best baseline classifier hyperparameters. The model’s performance is assessed with metrics such as AUC, F1-score, and Average Precision. Finally, SHAP analysis interprets predictions and identifies key features influencing patient satisfaction.

Our study focused on patient-specific clinical and demographic characteristics as potential predictors of treatment response, excluding measures of patient expectations that were the main focus of the original analysis^16^. The experimental protocol comprised questionnaires administered in German at three time points: on the day receiving the lumbar steroid injection, immediately prior to treatment, immediately after receiving the lumbar steroid injection and two weeks after the treatment.

To align with the study’s objective of evaluating predictors of treatment response, data collected immediately after the treatment were excluded. This exclusion was implemented to ensure a clear separation between the baseline data and the post-treatment data. The baseline was redefined as *T*_0_, representing the period prior to injection therapy, and the post-treatment period was labeled as *T*_1_, corresponding to two weeks after injection.

### Measures

Data collection encompassed a comprehensive set of questionnaire items addressing patients’ demographics, pain characteristics, and self-reported health status (Figure 1b).

### Demographics

Demographic information including age, sex, and education level, as well as categories of professional status — categorized as self-employed, student, homemaker, retired, incapacitated, or unemployed—was collected through questionnaires.

### Pain characteristics

The duration of back complaints was recorded into intervals, namely less than 4 weeks, 4 to 8 weeks, 8 to 12 weeks, and more than 12 weeks. Current back pain was assessed using numerical rating scales (NRS)^27^ which involve individuals rating their pain intensity on a scale from 0 (no pain) to 10 (worst pain imaginable). For participants who had previously undergone lumbar steroid injections, additional data were collected, including whether they experienced improvement after the last injection, the time elapsed since the previous treatment—categorized as less than 1 year, 1 to 2 years, or more than 2 years—and whether the procedure was performed by the same doctor or clinic. Motivation for treatment was assessed through sources of influence, including friends, family, the doctor performing the infiltration, general practitioner, internet, personal experience, and the importance of others’ opinions.

### Self-reported health status

A comprehensive set of validated PROMs covering medication beliefs, expectations, empathy in care, and self-efficacy were collected when infiltration therapy was administered (*T*_0_) to gain a multidimensional understanding of the patient’s pain experience and its impact. The items were extracted from widely used questionnaires in clinical research^26^ and consisted of the following: The Perceived Sensitivity to Medicine (PSM) scale was utilized to evaluate patients’ perceived responsiveness to medication in general^34^. This questionnaire includes items assessing perceived susceptibility to medications, beliefs about experiencing strong reactions, perceptions of having stronger reactions than others, and concerns about side effects from regular medication use. Responses were recorded on a 5-point Likert scale, ranging from “strongly disagree” to “strongly agree.” Furthermore, the Consultation and Relational Empathy (CARE) measure was employed to record their evaluation of the overall care experience^35^. This questionnaire assesses various aspects of the patient-provider interaction, such as the provider’s ability to make the patient feel at ease, allow them to tell their story, feeling understood by the healthcare provider, be interested in them as a whole person, fully understand their concerns, show care and compassion, and explain things clearly. Responses are given on a 5-point Likert scale ranging from “poor” to “excellent”. For multidimensional assessment of pain and disability, the Core Outcome Measures Index (COMI) back score was used^36,37^. The COMI questionnaire is a concise 7-item questionnaire which assesses the LBP disability, quality of life and pain perception including questions on back and leg pain intensity (0–10 NRS scale), function, symptom-specific well-being, general quality of life, and disability at work and social situations (5-point Likert scale)^36,38^. The COMI has been extensively validated^39–42^ against well established longer questionnaires such as the Roland Morris Disability Questionnaire^37,43^ and 36-item short-form health survey (SF-36)^44,45^. The Pain Self-Efficacy Questionnaire (PSEQ) was used to evaluate patients’ beliefs in their ability to cope with and manage pain despite its presence^46^. This questionnaire includes 10 items assessing the patient’s confidence in performing various activities despite pain, such as enjoying things, doing household chores, socializing, coping with pain without medication, achieving goals, engaging in leisure activities, coping with pain in general, accomplishing work tasks, leaving a normal lifestyle, and becoming more active. Responses are provided on a 7-point scale ranging from 0 (“not at all confident”) to 6 (“completely confident”).

### Outcomes

The primary focus of this study was to assess the effectiveness of lumbar steroid injections by evaluating both patient satisfaction and clinically meaningful improvements in pain intensity levels two weeks after treatment (*T*_1_). Accordingly, the main outcome was *pain level satisfaction*. A dichotomized variable was created based on the question: “Are you satisfied with the current pain level?” (“Sind Sie mit dem aktuellen Schmerzniveau zufrieden?”). Patients rated their satisfaction on a scale from 0 to 10, where 0 indicated complete satisfaction and 10 indicated no satisfaction at all. To align with clinical success criteria, a cut-off point of 6 was established. Patients scoring between 0 and 6 were classified as satisfied, reflecting a successful treatment outcome, while those scoring between 7 and 10 were classified as dissatisfied. Secondary outcomes focused on changes in pain intensity to objectively assess improvements in maximum pain levels. The baseline pain (*NRS*_*T*0_) level was computed as the maximum pain reported in the first two questions of the COMI questionnaire referring to back and leg pain evaluated using 0-10 NRS. The absolute change in pain (ΔPain) was calculated as the difference between baseline pain (*NRS*_*T*0_) and pain two weeks after treatment (*NRS*_*T*1_), computed as the maximum between back and leg pain reported at *T*_1_. To account for individual variability in baseline pain levels, a relative change in pain (Δ_*r*_Pain) was calculated by normalizing the absolute change to the baseline value, as follows:

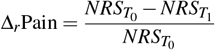

### Statistical Analysis

Figure 1c illustrates the comprehensive statistical analysis workflow, from initial data preprocessing through descriptive statistics to feature analysis of predictive models. Descriptive statistics were used to summarize the general characteristics of the participants across the two groups created according to the “*pain level satisfaction*” variable.

Continuous variables were reported as mean ± standard deviation. Categorical variables were presented as frequencies and percentages. For groups comparison, Student’s t-test (or Wilcoxon signed-rank tests when appropriate) and chi-square test were used for continuous and categorical variables, respectively. To control for multiple comparisons and maintain a false discovery rate of 5%, all statistical comparisons were adjusted using the Benjamini-Hochberg correction method. All statistical analysis were performed using the statsmodels library in Python version 3.10.

### Baseline predictive models of treatment satisfaction

A data-driven predictive model was developed to classify treatment outcomes based on the dichotomized pain level satisfaction variable, distinguishing between “satisfied” and “dissatisfied” patients. This section outlines the methodological approach used to design and benchmark predictive models. The development and implementation of the predictive models, were performed using Python version 3.10, with the scikit-learn and PyCaret^47^.

### Outer cross-validation data split

To mitigate the potential risk of overfitting associated with the limited sample size, a stratified nested cross-validation (CV) methodology was employed^48^. Nested CV involves two iteration loops over the data. In the first iteration, the outer loop applied a 10-fold stratified CV scheme to divide the dataset into training and testing sets, ensuring class balance (i.e., satisfied and dissatisfied) across folds and providing unbiased estimates of model generalization performance on completely unseen data.

### Feature engineering and data preprocessing

Prior to predictive model training, several preprocessing steps were performed to ensure the quality and relevance of the features. Features with missing values exceeding 15% were removed, given the limited dataset size, to prevent bias and ensure reliable analysis. The remaining missing data were imputed using an iterative approach: Random Forest (RF) was used for numeric features, and K-Nearest Neighbors (KNN) for categorical features, iterating five times for optimal imputation. Importantly, data imputation was completed prior to the outer cross-validation split to avoid any information leakage. Features were encoded based on their data type to ensure effective model training: numeric features were standardized using z-scores for consistent scaling, categorical variables were one-hot encoded to convert them into a numerical format, and ordinal features were label encoded to preserve their inherent order. To address potential multicollinearity and improve classification performance, features with a variance less than 0.01, as well as those with a correlation coefficient greater than 0.7, were removed.

### Baseline models training

During each outer loop cycle, the training data was further split using a 10-fold stratified inner CV to train the classifier. This step ensured the selection of optimal model configurations without introducing information leakage from the test set. This nested approach kept model training and hyperparameter tuning separate from the final performance evaluation, thereby improving the reliability of the generalization assessments.^49^.

Multiple baseline classifiers were trained on all the features and benchmarked using various classification algorithms: Logistic Regression^50^ (LR), KNN^51^, Support Vector Machines (SVM)^52^ with linear kernel^52^, Ridge Classifier^53^ (RC), Naive Bayes^54^ (NB), Linear (LDA) and Quadratic Discriminant Analysis^55^ (QDA), Decision Trees^56^ (DT), and ensemble methods including Extra Trees^57^ (ET), RF^58^, AdaBoost^59^, Gradient Boosting Machine^60^ (GBM), XGBoost^61^, and LightGBM^62^. These baseline comparison provided a reference point for subsequent optimization.

### Classification model performance evaluation

Predictions from all outer loop iterations were concatenated to calculate the final performance metrics, providing a robust estimate of the model’s generalization ability while maintaining strict train-test separation. Model performance was primary assessed based on F1-score, average precision (AP), and Matthews correlation coefficient (MCC), which are classification metrics particularly useful for imbalanced datasets. The F1-score represents the harmonic mean of precision and recall calculated as:

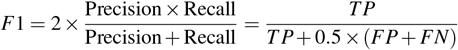

where TP, FP, and FN denote true positives, false positives, and false negatives, respectively. The F1 score ranges from 0 to 1, with 1 indicating perfect precision and recall, and 0.5 indicating random guessing. The AP for a given class is calculated as the area under the precision recall curve:

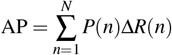

where *P*(*n*) denotes precision at the *n*-th recall level, while Δ*R*(*n*) measures changes between consecutive recall levels. AP ranges from 0 to 1, with 1 indicating optimal precision and recall and 0 indicating performance equivalent to random guessing. MCC is a robust summary metric computed as the correlation coefficient between observed and predicted binary classifications^63^:

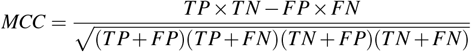

MCC values range from -1 to +1, where +1 represents a perfect prediction, 0 indicates that the prediction is not better than a random prediction, and -1 represents total disagreement between prediction and observation. In addition, standard classification metrics including precision, recall, accuracy, Cohen’s Kappa coefficients, and Area under the Receiver Operating curve (AUC) were used for a comprehensive evaluation.

### Optimized predictive models of treatment satisfaction

Building on the baseline comparison, the best-performing model from the cross-validation splits was further tuned (Figure 1d) with the objective of identifying the most informative clinical and demographic characteristics through statistical techniques. The data encoding procedures, i.e., encoding and standardization of variables, remained consistent with the approach used in the baseline classifiers, ensuring methodological uniformity.

### Feature selection and oversampling

Next, feature selection was performed according to an XGBoost estimator based importance ranking, retaining the top 70% (35 features). To address the class imbalance in the dataset, random oversampling was applied during model training. This technique involved duplicating samples from the minority class to balance the class distribution.

### Optimized model training and hyperparameter optimization

We optimized hyperparameters for the best-performing baseline model, RF, using Bayesian and random grid search. This optimization aimed to identify the most informative clinical and demographic characteristics through systematic exploration of the model’s parameter space while minimizing overfitting. The hyperparameters were tuned through 5-fold stratified cross-validation (inner folds) for each outer fold, with predefined hyperparameter space ranges.

The RF model was configured with 10 to 1000 trees, with a higher number of trees that potentially improve performance, but increase computational time. The tree depth ranged from 1 to 32, allowing the model to capture more complex patterns, although deeper trees carry a higher risk of overfitting. The risk of overfitting was mitigated by adjusting split node samples (2 to 20) and leaf samples (1 to 20). The feature fractions for splitting were varied between 0.1 and 1.0, with smaller values introducing randomness to reduce overfitting.

### Interpretability

Feature importance was assessed using SHapley Additive exPlanations (SHAP), which identified the principal predictors for the best-performing classifier. SHAP values provide a quantitative measure of the influence of individual features, representing the average marginal contribution of each feature to the model’s prediction for a given instance^64^. These values are computed by comparing the model’s predictions with and without each feature, considering all possible feature combinations^64^. Larger absolute SHAP values indicate stronger effects, while the sign of the value shows whether a feature with a positive SHAP value increases or decreases the prediction outcome.

### Clinical important absolute and relative change in patient-reported pain intensity

ROC analysis (Figure 2) was performed to identify meaningful thresholds for both absolute (ΔPain) and relative (Δ_*r*_Pain) changes in patient-reported pain scores reduction after lumbar infiltration, using ‘pain level satisfaction’ as a reference variable. The ideal point on an ROC curve^65^, would be in the upper left corner (0, 1), representing the best trade-off between specificity (Sp) and sensitivity (Se) for a diagnostic test (Sp 100%, Se 100%)^66^. In our analysis, the optimal cut-off points on these curves would represent the smallest change in pain score, absolute and relative, that best distinguishes between satisfied and unsatisfied patients. The optimal cut-off point can be determined using several approaches^67^.

- **The Euclidean distance method**: Finds the ROC point closest to the ideal cut-off (0,1) by minimizing the Euclidean distance^67^.
- **Youden method**: The Youden Index^68^ measures diagnostic performance by summing sensitivity (Se) and specificity (Sp): J = Se + Sp − 1 = Se − (1 − Sp). The optimal cut-off is at the maximum Youden Index, with higher values indicating greater effectiveness by maximizing the vertical distance from the random ROC (diagonal line).
- **Farrar method**: The Farrar method determines the threshold at which sensitivity and specificity are equal^67^, to equally identify satisfied (Se) and unsatisfied patients (Sp).

**Figure 2.**
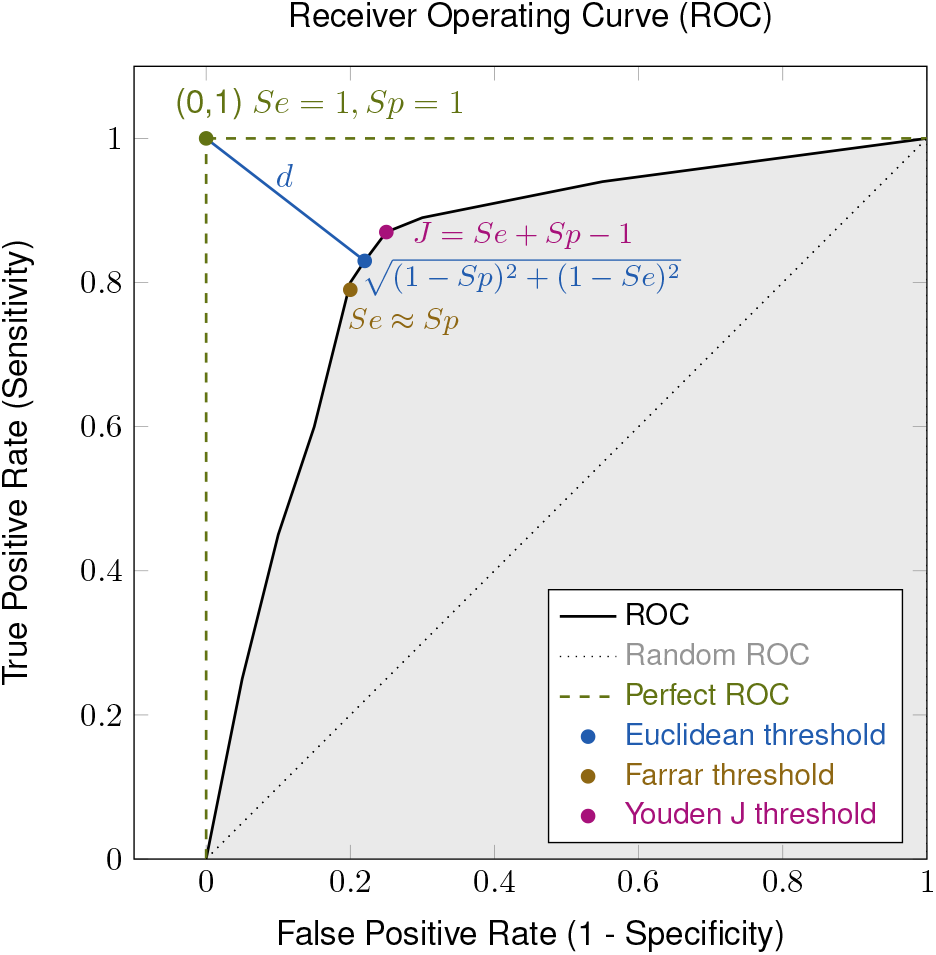
The Receiver Operating Characteristic (ROC) curve provides an assessment of the predictor’s efficacy by plotting True Positive Rate or Sensitivity (Se) against False Positive Rate, or 1-Specificity across multiple binarization thresholds. The ideal ROC curve (dashed green), indicative the ideal classification, and the diagonal (dotted gray) representing random prediction are included for comparative purposes. The highlighted (gray) Area Under the Curve (AUC) serves as a summary statistic for overall classifier performance. The methods employed for determining a threshold that optimally equilibrates sensitivity and specificity comprise: the Euclidean Distance approach (blue), involving the minimization of distance (*d*) to the ideal threshold at left corner (0, 1); Youden’s Index, which focuses on maximizing the disparity between the *Se* + *Sp* − 1, finding the point furthest from the diagonal ROC; the Farrar Method (bronze), where the Sp equals Se, representing a balance between false positives and the complementary of false negatives.

### Predictor of treatment success based on change in patient-reported pain intensity

To further assess the robustness of our classification approach under a clinically meaningful definition of success, we conducted an additional evaluation using the smallest relative change in baseline pain scores (Δ_*r*_Pain) as a threshold. Specifically, based on the optimal cut-off identified in our ROC analysis, we stratified participants into “satisfied” or “dissatisfied” groups according to whether their relative reduction in self-reported pain exceeded this threshold at *T*_1_. This aims to reflect a more clinically relevant perspective of improvement, since percentage-based reductions in pain often better capture individual differences than absolute changes alone.

In this simplified analysis, we applied the same preprocessing and cross-validation schemes but focused exclusively on the set of baseline classifiers without hyperparameter optimization (see previous section). By benchmarking these models, we obtained a clear view of how different algorithms perform when using a clinically significant cut-off for pain relief, rather than the original dichotomous satisfaction variable.

## Results

### Demographics and baseline pain characteristics

Table 1 presents the sociodemographic characteristics of 212 patients stratified by pain level satisfaction, with 158 patients (75%) in the satisfied group and 54 patients (25%) in the dissatisfied group, with the mean age being 67.8 years (SD = 2.7), or gender distribution, with 53% of the patients being female. The analysis did not reveal statistically significant differences in age and gender distribution for the 2 groups. Educational backgrounds were comparable between groups, with vocational apprenticeships being the most common qualification, followed by higher vocational education. Professional status was similarly distributed across groups, with nearly half of the participants in both groups being retired. No statistically significant differences were observed between the groups in terms of education or employment status.

**Table 1.** Descriptive statistics of sociodemographic variables stratified by reported pain level satisfaction. Data are presented as mean (± standard deviation) for continuous variables and frequency (percentage) for categorical variables. Statistical comparisons p-values are corrected via Benjamini-Hochberg method

|  | Satisfied (n=158) | Dissatisfied (n=54) | p-value<sub>fh</sub> |
| --- | --- | --- | --- |
| **Demographics** |  |  |  |
| Age, mean (SD) | 67.8 (±12.7) | 63.0 (±16.4) | 0.39 |
| Sex - female, N (%) | 83 (52.5) | 33 (61.6) | 0.72 |
| **Worktime** |  |  |  |
| Unknown, N (%) | 116 (73.4) | 33 (61.1) | 0.43 |
| Full time, N (%) | 25 (15.8) | 13 (24.1) | 0.59 |
| Part time, N (%) | 17 (10.8) | 8 (14.8) | 1.00 |
| **Education** |  |  |  |
| Unknown, N (%) | 7 (4.4) | 2 (3.7) | 1.00 |
| No school, N (%) | 0 (0.0) | 1 (1.9) | 1.00 |
| Secondary School, N (%) | 9 (5.7) | 2 (3.7) | 1.00 |
| Vocational Apprenticeship, N (%) | 58 (36.7) | 22 (40.7) | 1.00 |
| Vocational-Professional Baccalaureate, N (%) | 9 (5.7) | 3 (5.6) | 1.00 |
| Academic Baccalaureate, N (%) | 6 (3.8) | 1 (1.9) | 1.00 |
| Higher vocational education, N (%) | 32 (20.3) | 11 (20.4) | 1.00 |
| University Degree, N (%) | 27 (17.1) | 9 (16.7) | 1.00 |
| Doctorate, N (%) | 10 (6.3) | 3 (5.6) | 1.00 |
| **Profession (not exclusive)** |  |  |  |
| Self-employed, N (%) | 28 (17.7) | 4 (7.4) | 0.42 |
| Student, N (%) | 2 (1.3) | 1 (1.9) | 1.00 |
| Housework, N (%) | 32 (20.3) | 3 (5.6) | 0.33 |
| Retired, N (%) | 80 (50.6) | 21 (38.9) | 0.51 |
| Incapacity, N (%) | 9 (5.7) | 8 (14.8) | 0.39 |
| Unemployed, N (%) | 1 (0.6) | 0 (0.0) | 1.00 |
| Profession: other, N (%) | 2 (1.3) | 1 (1.9) | 1.00 |

The majority of patients reported significant baseline pain, with a mean pain level of 5.8 *±* 2.3 on the NRS (0-10), indicating substantial discomfort before treatment. Notably, 75% of patients had baseline pain levels of 5 or higher. After treatment, patients experienced a reduction in pain, with the mean pain level decreasing to 3.9 ± 2.6, reflecting an average pain reduction of 2.7 ± 2.5 points on the NRS. At baseline (*T*_0_), dissatisfied patients reported maximum higher back or leg pain levels compared to satisfied patients, 6.4 *±* 2.1 vs. 7.3 *±* 2.4 (*p* = 4.865*e* − 03). After treatment (*T*_1_), this difference became more pronounced, with satisfied patients showing lower pain scores of 3.1 *±* 2.2 while dissatisfied patients maintained high pain levels (6.2 *±* 1.9, *p* = 5.685*e*− 17)

### Baseline predictive models of treatment satisfaction

We evaluated baseline classifiers with stratified cross-validation and quantified performance using per-fold mean AP, AUC, accuracy, F1-score, Cohen’s Kappa, and MCC without hyperparameter optimization, summarized in Figure 3.

**Figure 3.**
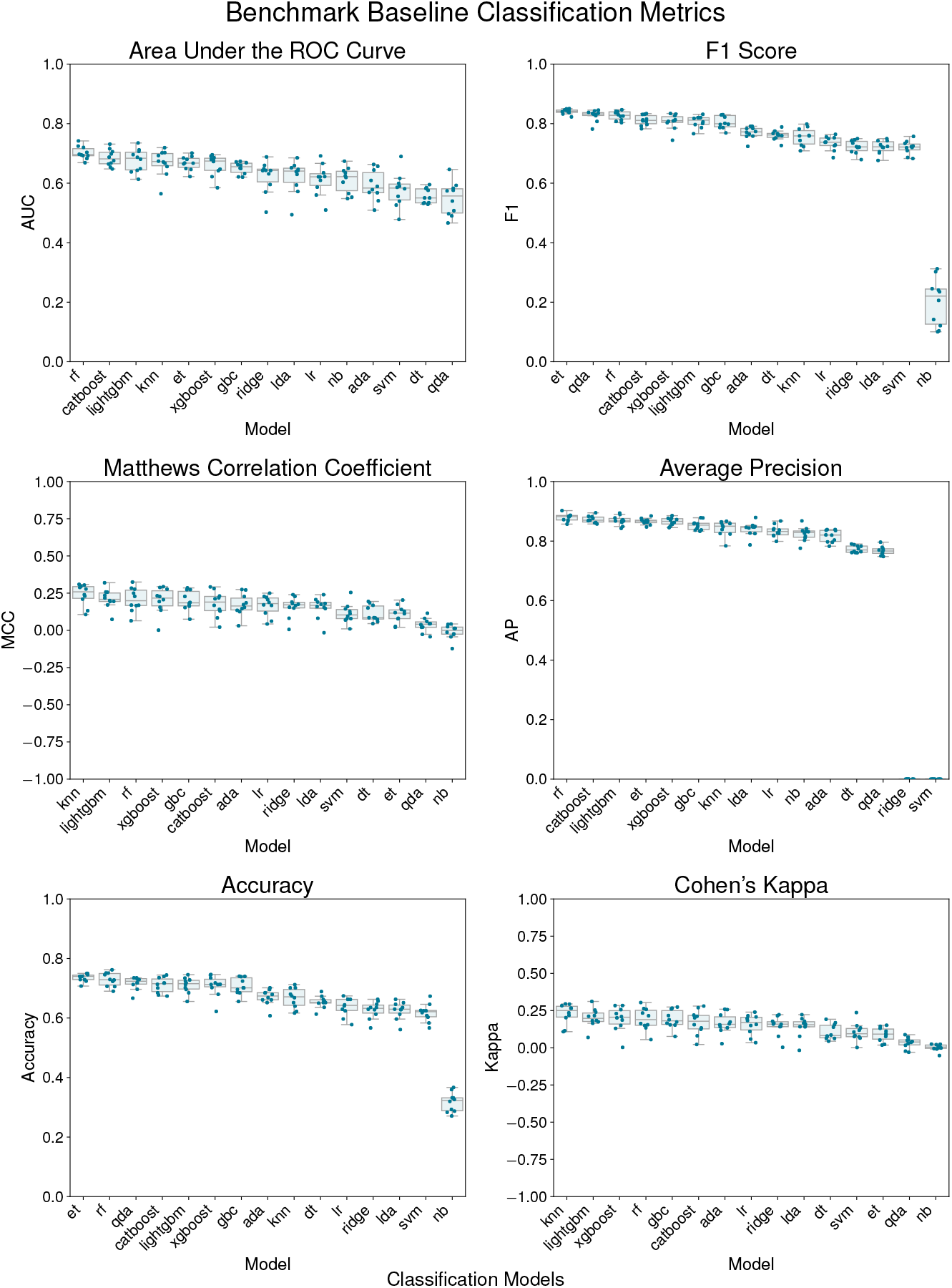
The boxplots illustrate the distribution of each metric across cross-validation folds for each classifier model, facilitating a performance comparison of baseline classifiers utilizing various metrics: Area Under the ROC Curve (AUC), F1-score, Matthews Correlation Coefficient (MCC), Average Precision (AP), Accuracy, and Cohen’s Kappa. The classifiers included K-Nearest Neighbors (KNN), Random Forest (RF), Extra Trees (ET), Gradient Boosting Classifier (GBC), XGBoost, Ridge Classifier (RC), Linear Discriminant Analysis (LDA), LightGBM, Logistic Regression (LR), Decision Tree (DT), Support Vector Machine (SVM), AdaBoost, Naive Bayes (NB), and Quadratic Discriminant Analysis (QDA). RF achieved the highest mean Average Precision (AP) of 0.879 *±* 0.012 and an Area Under the Curve (AUC) of 0.702 *±* 0.020. RF also exhibited strong classification accuracy, with a mean of 0.729 *±* 0.022 and an F1-score of 0.827 *±* 0.014. In contrast, K-Nearest Neighbors (KNN) achieved the highest Cohen’s Kappa value of 0.228 *±* 0.065 and the highest Matthews Correlation Coefficient (MCC) value of 0.239 *±* 0.068.

The RF demonstrated superior performance in terms of discriminative ability, achieving the highest AP of 0.856 on the full cross-validated test set. In terms of overall metrics, the model showed an accuracy of 70.3%, a precision of 78.1%, and a recall of 83.5%, resulting in an F1-score of 0.807. The MCC and Cohen’s Kappa scores were 0.163 and 0.161, respectively, suggesting a weak correlation and fair agreement between predicted and actual classes. The ROC AUC was 0.731, indicating fair discriminative ability.

### Optimized classifier performance

Although the optimal hyperparameters for the RF classifier exhibited slight variations across different folds, certain consistent trends were discernible. The number of estimators demonstrated a wide range, ranging from 21 to 109 trees. Tree depth varied, with some folds using shallow trees (depths of 4-6) and others can grow without a predetermined limit. Feature sampling consistently targeted 70-90% of features. Conservative parameters for leaf nodes and node splitting required minimal samples (2-5 for leaves, 6-10 for nodes).

The tuned RF classifier demonstrated moderate predictive performance across multiple evaluation metrics, as visualized in Figure 4a. The model achieved a ROC AUC of 0.688, indicating fair discriminative ability. The Precision-Recall curve (right of Figure 4a) revealed an AP score of 0.846, showcasing a good precision-recall trade-off. The confusion matrix (Figure 4a) shows the model correctly identified 131 satisfied and 27 dissatisfied patients, while misclassifying 29 as satisfied and 25 as dissatisfied out of 212 patients. This distribution highlights a slight class imbalance, with the model demonstrating higher sensitivity for satisfied patients.

**Figure 4.**
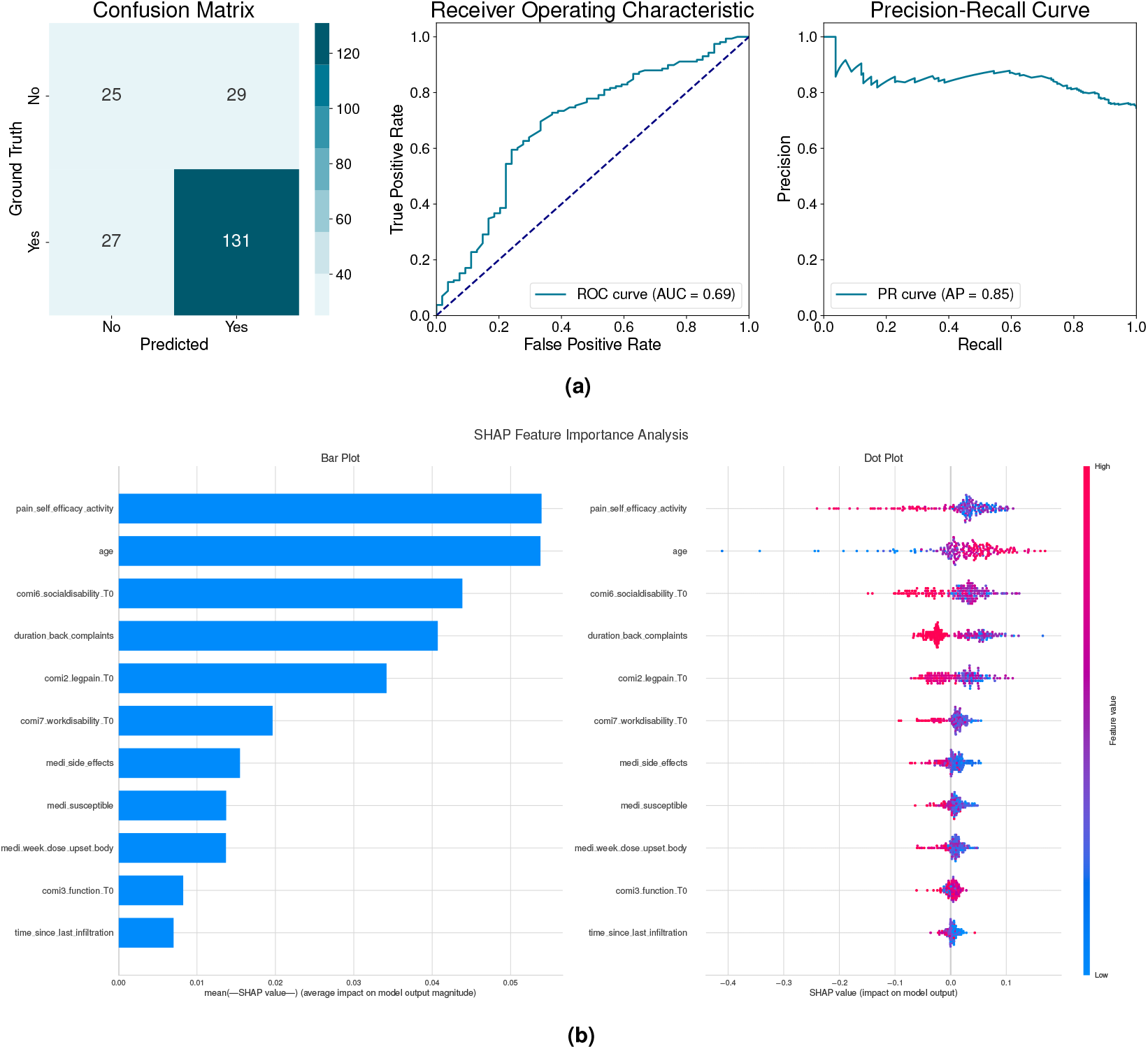
(a) **Classification Results of the optimized Random Forest (RF) model for patient reported pain level satisfaction two weeks after lumbar steroid injection treatment** (left) The Confusion Matrix shows the true and false positives and negatives classification (middle) Receiver Operating Curve (ROC) curve (right) Precision-Recall Curve illustrating he relationship between precision and recall across classification thresholds. **(b) SHAP Feature Importance Analysis Plots**: Bar plot (left) illustrate the average relative significance and dot plots (right) reveals how each feature affects outcomes, where red dots indicate higher values and blue dots show lower values. The bar plot underscores the influence of pain self-efficacy activity, age and disability as well as pain temporal variables in the model outcome.

The F1 score of 0.824 reflected balanced performance between precision (0.819) and recall (0.829). Further evaluation of performance metrics provided additional insights: the MCC and Cohen’s Kappa values of 0.296 indicated weak correlation and fair agreement between predicted and actual classes. The balanced accuracy of 64.6% further underscored the model’s moderate classification capabilities, with an overall accuracy of 73.6%.

### Feature importance analysis of optimized classifier

The SHAP analysis of the common features selected across all folds revealed that pain self-efficacy features were the most significant predictors of post-treatment pain level satisfaction, as illustrated in the bar plot in Figure 4b.

The strongest predictor was patients’ perceived ability to stay active despite pain (*pain_self_efficacy_activity*), followed by patient age and days of activity limitation due to back pain(*comi6_socialdisability_T0*). The duration of severe back symptoms (*duration_back_complaints*) and baseline leg pain intensity *(comi2_legpain_T0)* showed moderate predictive influence. Medication sensitivity factors including history of side effects *(medi_side_effects)*, self-reported susceptibility *(medi_susceptible)*, and reaction to low doses *(medi_week_dose_upset_body)* exhibited weaker but statistically meaningful associations. Work disability days *(comi7_workdisability_T0)* and baseline functional impairment *(comi3_function_T0)* completed the predictor incluence ranking.

### Clinical important change in patient-reported pain intensity and outcomes

The results in Figure 5 present optimal thresholds for absolute and relative changes in pain scores from ROC curve analysis to distinguish between satisfied and dissatisfied patients after steroid injection therapy.

**Figure 5.**
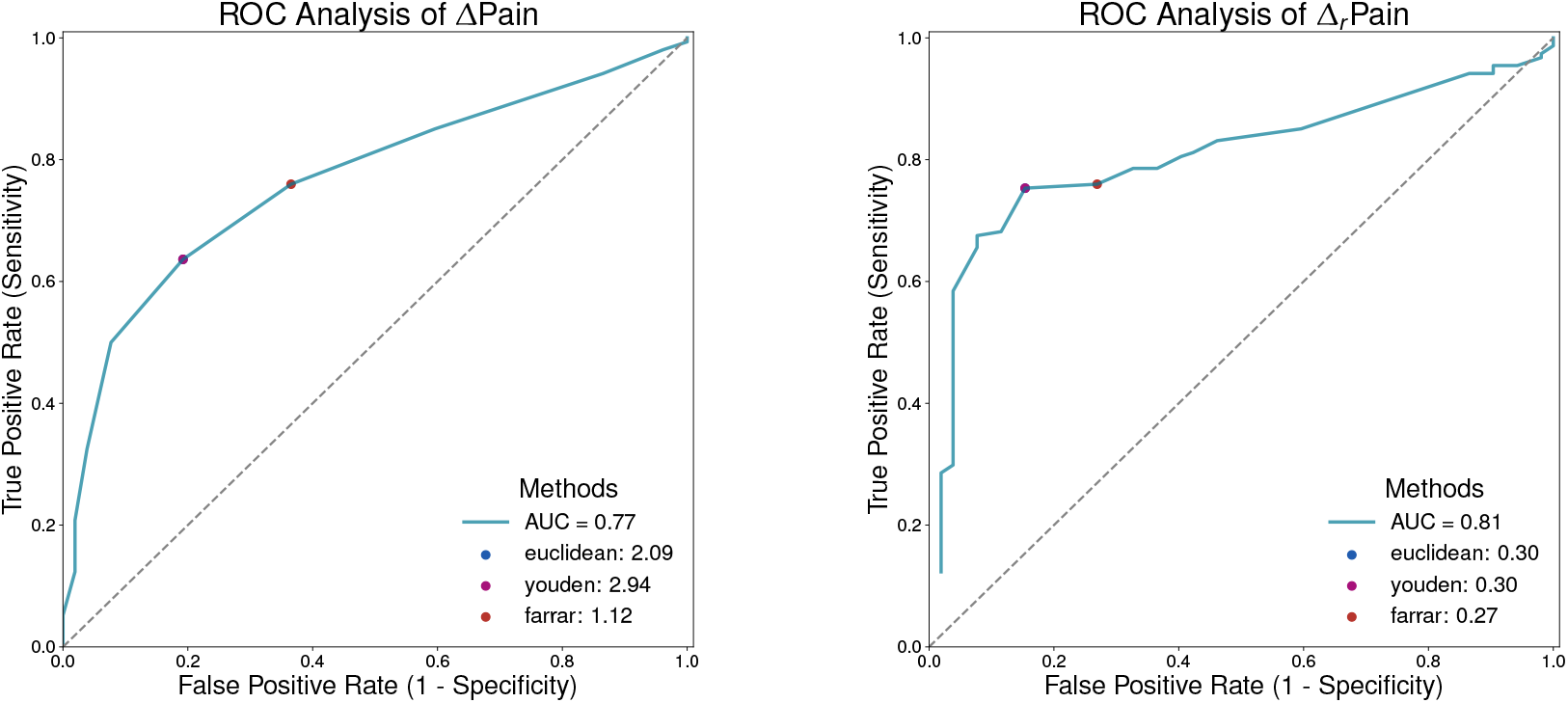
ROC Curve Analysis for absolute pain level (Δ*Pain*) and relative (Δ_*r*_*Pain*) change in reported maximum pain between baseline and two-weeks after treatment. The figure illustrates the best thresholds of both changes in pain scores that can be used in distinguishing between satisfied and dissatisfied patients following lumbar steroid injection therapy.

The ΔPain threshold achieved an AUC of 0.77, while the Δ_*r*_Pain cut-off point had an AUC of 0.81, indicating a strong potential to discriminate between satisfied and dissatisfied patients.

For the absolute change in pain ΔPain, the Euclidean distance method identified an optimal threshold of 2.09 (sensitivity: 0.64, specificity: 0.81), while the Youden index yielded a higher threshold of 2.94 with identical sensitivity (0.64) and specificity (0.81). The Farrar method suggested a lower threshold of 1.12, achieving higher sensitivity (0.76) but lower specificity (0.63).

For the absolute change in pain (ΔPain), the Euclidean distance method determined a threshold of 2.09, characterized by a sensitivity of 0.64 and a specificity of 0.81. Meanwhile, the Youden method identified a threshold of 2.94 accompanied by a sensitivity of 0.64 and a specificity of 0.81. The Farrar method yielded a threshold of 1.12, with a sensitivity of 0.76 and a specificity of 0.63. Considering the relative change in pain (Δ_*r*_pain), both the Euclidean distance and Youden methods determined an optimal cut-off point of 0.30, with a sensitivity of 0.75 and a specificity of 0.85. The Farrar method identified a threshold of 0.27, with a sensitivity of 0.76 and a specificity of 0.73.

Taking into account the consistency between methods and the balance between sensitivity and specificity, a ΔPain of 2.09 and a Δ_*r*_Pain of 0.30 were identified as the best thresholds indicating the minimal clinically relevant enhancement in pain relief after steroid injection therapy.

### Baseline classifiers for treatment success based on change in patient-reported pain intensity

A re-assessment of the baseline classifiers was conducted to predict patient satisfaction based on the previously established clinically meaningful threshold of 30% relative pain reduction (Δ_*r*_Pain = 0.30). RF demonstrated superior performance in terms of discriminative ability, achieving the highest mean AP of 0.757*±*0.020 and AUC of 0.651*±*0.020. Extra Trees (ET) exhibited strong performance with a mean AUC of 0.628*±*0.024 and accuracy of 0.579*±*0.020 for relative pain reduction. MCC values were consistently low across all models, ranging from 0.003 for QDA to 0.196 for RF indicating weak correlation between predicted and actual classes. Similarly, Cohen’s Kappa values were also modest, with the highest value being 0.190 for RF, followed by 0.172 for AdaBoost.

## Discussion

Our findings underscore the complex and multifaceted nature of CLBP and highlight the potential of data-driven predictive modeling to refine therapeutic strategies for lumbar steroid injection therapy. The results demonstrate that baseline characteristics, particularly psychosocial factors such as pain self-efficacy, play a crucial role in determining patient satisfaction after treatment. Previous research showed the importance of expectation on treatment outcome^16^. These insights provide a strong foundation for the development of personalized treatment plans aimed at optimizing outcomes in CLBP patients.

The predictive model developed in this study represents a meaningful advance in understanding and addressing the heterogeneity of treatment responses. By highlighting individualized predictors like pain self-efficacy, it enables personalized treatment strategies and better resource allocation. Additionally, the model’s use of outcome-driven metrics, such as clinically meaningful pain reduction thresholds, provides actionable insights for standardizing evaluations and optimizing patient-centered care in CLBP.

Using data-driven machine learning algorithms and rigorous cross-validation, the model achieved moderate performance, with an F1 score of 0.812 and an area under the precision-recall curve of 0.858 with an F1 score of 0.824 and an area under the precision-recall curve of 0.846. These metrics underscore the model’s capacity to identify patients most likely to benefit from lumbar steroid injections, offering a practical tool for clinicians to enhance treatment allocation. However, the relatively modest Matthews Correlation Coefficient (MCC: 0.296) and Cohen’s Kappa (0.296) indicate the need for further refinement to improve discriminatory power and reduce misclassification. The divergence between strong discriminative ability and weaker correlation metrics reflects the inherent complexity of predicting treatment success in CLBP patients based on patient-reported outcomes.

The feature importance analysis underscores the multidimensional nature of CLBP outcomes, with psychosocial factors emerging as the most significant predictors of patient satisfaction. Specifically, SHAP analysis revealed that *pain self-efficacy activity*—the ability to stay active despite pain—was the strongest contributor to post-treatment satisfaction. This finding highlights the critical role of psychological resilience in shaping patient outcomes, aligning with prior research emphasizing the importance of self-efficacy in chronic pain management^69^

Demographic and functional variables emerged as significant contributors to treatment outcomes in CLBP. Patient age and days of activity limitation due to back pain (*comi6_socialdisability_T0*) ranked as the second and third most influential predictors, respectively. These findings suggest that older patients and those with greater activity limitations may experience worse treatment outcomes, consistent with prior research highlighting the impact of age and functional capacity on recovery trajectories in CLBP^70^.

Clinical factors, including the duration of severe back symptoms (*duration_back_complaints*) and baseline leg pain intensity (*comi2_legpain_T0*), also demonstrated moderate influence. These variables reflect the complex interplay between symptom severity and treatment response, as supported by studies identifying symptom duration and baseline pain intensity as important prognostic factors for pain reduction and disability improvement in multidisciplinary treatment programs for LBP^71,72^.

The ROC analysis revealed the minimum score for pain reduction that patients perceive as beneficial after steroid injection therapy. The relative change in pain intensity demonstrated superior discriminative capabilities based on the ROC-AUC analysis (Figure 5). The AUC for the relative change threshold was slightly higher (0.81 vs 0.78) than that for the absolute change threshold, suggesting that considering the percentage reduction in pain might be more accurate in predicting patient satisfaction than the absolute change. This superiority can be explained by the percentage reduction’s ability to account for baseline variability in pain intensity. Patients with higher baseline pain levels may require larger absolute reductions to perceive meaningful relief, while those with lower baseline levels may find smaller absolute reductions sufficient. By normalizing pain reduction relative to the initial intensity, percentage change offers a more individualized and context-sensitive measure, better capturing patient satisfaction across a diverse population. The identified pain reduction thresholds, consisting of an absolute change of 2.0 NRS points and a relative reduction of 30%, are consistent with the range of reductions previously documented in patients with acute pain^28^. These thresholds provide objective measures for assessing treatment outcomes, managing patient expectations, and standardizing CLBP steroid injection evaluations in clinical settings.

## Limitations

Despite its strengths, our study has several limitations. First, the sample size was relatively small, which may have limited the ability of the model to generalize to broader populations. Furthermore, the use of PROM as the primary endpoint, while clinically relevant, is subjective in nature and may be influenced by recall bias or patient expectations. Furthermore, the dichotomization of the satisfaction outcome variable may oversimplify the complexity of pain level satisfaction. Although this binary classification is necessary for analysis, it can potentially lead to a loss of granularity with respect to degrees of satisfaction and their contributing predictors. Future models should employ ordinal or continuous outcomes for deeper insights into patient satisfaction and treatment response. Exploring the integration of longitudinal data, including repeated assessments of pain and function, may also provide a more dynamic understanding of treatment responses. Future research should validate these findings in larger and more diverse cohorts and incorporate objective measures, such as functional imaging or biomarker analysis, to complement self-reported data.

Future research should aim to enhance the data-driven predictive model by incorporating additional relevant factors, with a focus on multimodal variables that better capture the heterogeneity of CLBP. Integrating imaging data and more comprehensive psychosocial assessments could significantly improve the model’s predictive accuracy and clinical applicability. Prospective studies with larger, more diverse cohorts and extended follow-up periods are essential to validate and build upon these findings. Additionally, examining the utility of identified thresholds for relative and absolute changes in pain intensity across distinct phenotypes of CLBP may offer valuable insights for optimizing conservative treatment strategies.

## Conclusion

In summary, this study underscores the intricate nature of CLBP and the potential of predictive modeling to inform more personalized treatment approaches. Psychosocial factors, particularly pain self-efficacy, emerged as significant contributors to patient satisfaction, reinforcing the need to address psychological dimensions alongside physical interventions. The identified thresholds for pain reduction provide practical benchmarks for evaluating treatment outcomes and standardizing clinical practices. Despite promising predictive performance, the model’s limitations highlight the necessity for further refinement and validation with larger, more diverse populations. Future efforts should prioritize integrating multimodal data, such as imaging and comprehensive psychosocial assessments, to enhance the predictive power and clinical utility of these models.

## Data availability statement

Anonymized data used in this study will be made available upon reasonable request to and in compliance with the General Data Protection Regulation (EU GDPR). We publish all code required to reproduce the presented results in our GitLab repository: https://gitlab.ethz.ch/BMDSlab/publications/low-back/infiltration-outcome-pain-prediction

## Acknowledgements

This project was supported by grant (# 380, Jutzeler, Manjaly) of the Strategic Focus Area “Personalized Health and Related Technologies (PHRT)” of the ETH Domain (Swiss Federal Institute of Technology). We sincerely thank the patients who participated in this study, contributing their time and experiences to enhance our understanding of chronic low back pain treatment. We also acknowledge and thank the data collection team at Schulthess clinic, as well as the researchers and clinicians of the initial TREXI study^16^. ZMM is grateful for support by the Wilhelm-Schulthess Stiftung.

We also gratefully acknowledge the support of the Language Center of UZH and ETH Zurich, with special thanks to Kimberly Lewis for her expert assistance in improving the clarity and quality of this manuscript’s scientific writing. For the development of this work, AI-assisted coding systems such as Copilot and Perplexity were used. During the preparation of this manuscript, the author(s) reviewed the text for grammar correctness and enhanced the content with the assistance of AI generative tools (Writefull and Perplexity). After using this tool/service, the author(s) reviewed and edited the content as needed and take(s) full responsibility for the content of the publication. The figure 1 has been designed using resources from Flaticon.com.

## Author contributions statement

M.M. conceptualized the study, developed the codebase, conducted the experiments, and drafted the manuscript. I.D.S. contributed to data preprocessing, performed statistical analysis, and assisted with data interpretation. A.C. provided support with the ROC analysis and participated in the design of the experiments. R.E. contributed to data collection and offered clinical expertise. F.G. provided clinical expertise and assisted in the design of the experiments. C.R.J. and Z.M.M. supervised the project, offered critical revisions, and ensured the scientific rigor of the study. All authors critically reviewed and approved the final version of the manuscript.

## Additional information

C.R.J. serves as a scientific consultant to Abbvie and Mitsubishi Takeda; however, this role had no influence on the design, conduct, or reporting of this study. All other authors declare no conflict of interest.

